# Assessing Patient Safety Culture Among Doctors, Nurses, and Allied Health Professionals Using the Indian Version of the Hospital Survey on Patient Safety Culture (I-HSOPSC 2.0): A Cross-Sectional Study at a Tertiary Care Hospital in India

**DOI:** 10.64898/2026.09.15.26363107

**Authors:** T M Ridha Sulaiha, Kavitha Ashok, Aqueel Fatma Syed

## Abstract

**Background:** A culture of patient safety is a significant determinant of healthcare quality and the prevention of patient harm. The objective of this study was to evaluate patient safety culture among healthcare workers at a tertiary-care hospital in India using the Indian-adapted Hospital Survey on Patient Safety Culture (I-HSOPSC 2.0).

**Method:** The survey was descriptive and cross-sectional, carried out among 96 healthcare practitioners, comprising 32 doctors, 32 nurses, and 32 allied health professionals. Also, a validated version of the Hospital Survey on Patient Safety Culture (I-HSOPSC 2.0) was used to measure patient safety culture in India. Descriptive statistics, Shapiro-Wilk test, Kruskal-Wallis test, Fisher’s exact test, and positive response rate analysis were used for the data analysis.

**Result:** The overall patient safety culture score was 3.72 ± 0.52, indicating a positive view of patient safety culture. In terms of domains, Communication about Errors, Management Support for Patient Safety, and Teamwork scored highest, while Staffing, Work Pace, and Response to Errors scored lowest. Considerable discrepancies were observed across professions, with allied health professionals giving more favourable answers than nurses.

**Conclusion:** Overall, the culture around patient safety was good, but areas such as staffing, pace of work, and error response need targeted attention to sustain patient safety and improve the quality of care in healthcare services.

## Introduction

Hospitals are crucial in delivering preventive, diagnostic, therapeutic, and rehabilitative health care services. Moreover, as health care becomes more complex, a committed patient safety culture is vital to maintaining service quality. Patient safety culture encompasses the shared beliefs, values, and behaviours of health care providers that prioritise safety through effective communication, cooperation, and error avoidance. Hence, a positive safety culture leads to sound clinical decisions, reducing adverse incidents and improving health outcomes. The safety culture of patients is tied to workplace variables such as aggression, fatigue, and job satisfaction, all of which contribute to good patient care practices and patient safety. Each year, millions of avoidable adverse incidents occur. These incidents are mostly the result of the organisation’s failures rather than of mistakes committed by individuals. A strong safety culture fosters all types of incident reporting, good communication, learning within the organisation, openness, and a non-punitive attitude toward employee mistakes [1–4].

While patient safety is a critical aspect of healthcare, little is known about patient safety culture in developing countries like India. Safety culture encompasses the shared attitudes, beliefs, and values that influence how individuals approach safety within an organisation. Hospitals face numerous challenges, including error management, incident reporting, and the delivery of quality care. Patient harm can lead to the closure of public hospitals. In hospitals, various tools have been created to measure patient safety culture; among the most widely accepted and validated are the Safety Attitudes Questionnaire (SAQ) and the Hospital Survey on Patient Safety Culture (HSOPSC). The latter tool, developed by the Agency for Healthcare Research and Quality (AHRQ), is widely adopted to measure safety improvement projects. The modified HSOPSC 2.0, implemented in 2019 and comprising 32 items across 10 areas of safety culture, represents a holistic approach to assessing safety culture in healthcare facilities [5–7].

The Indian Hospital Survey on Patient Safety Culture 2.0 (I-HSOPSC 2.0) is an approved measure adapted for Indian healthcare professionals to assess their perceptions of patient safety culture in India. This measure comprises 32 core items across 10 aspects of patient safety culture, covering issues such as teamwork, error-related communication, management support for patient safety, staffing and work pace, error response, organisational learning, and patient safety incident reporting. Thanks to the adoption of this universal measure, validated for India, it is possible to conduct a systematic assessment of healthcare professionals’ views on patient safety culture. The instrument identifies strengths and weaknesses in patient safety within healthcare organisations. It has demonstrated good psychometric properties in the Indian context [8–10].

While patient safety culture is increasingly studied in hospitals, there remains a need for more evidence from diverse healthcare settings across India. The Indian validation of I-HSOPSC 2.0 has demonstrated its reliability and effectiveness in evaluating patient safety culture among professionals, but additional studies are needed across diverse medical settings and institutions [1]. Studies in India have recently assessed patient safety culture using the HSOPSC 2.0 across various tertiary care settings and among healthcare professional groups, including doctors, nurses, and allied health professionals [11]. In addition, other studies in India have investigated patient safety culture among hospital nurses, focusing on HSOPSC 2.0 [12]. A recent study conducted at a public tertiary-care teaching hospital in Central India examined patient safety culture among various professional groups, including physicians, nurses, and allied healthcare professionals [13]. Other studies have indicated differences in patient safety culture across healthcare institutions and their governing characteristics; thus, the local organisational context should be evaluated in this area.

Also, assessing patient safety culture at the level of the hospital organisation can reveal strengths and areas for change and help healthcare managers understand how to develop appropriate patient safety-related initiatives. Recent evidence from Bangladesh similarly demonstrates the use and cultural adaptation of HSOPSC 2.0 in an LMIC context. However, the study was limited to nurses, indicating a need for further research involving physicians and other healthcare professionals [14].

Even with the increasing adoption of HSOPSC 2.0 in healthcare facilities in India and other LMIC countries, there isn’t enough information available to compare patient safety culture among doctors, nurses, and other healthcare workers within the same hospital. This is important because the various roles, duties and working conditions of the professionals can affect their views on patient safety culture. Hence, the study was conducted to measure and compare patient safety culture among doctors, nurses, and other health professionals using the I-HSOPSC 2.0.

### Objectives

1. To assess patient safety culture among doctors, nurses and allied health professionals using the I-HSOPSC 2.0 tool
2. To evaluate the frequency of patient safety event reporting among these professional groups.
3. Compare the item-level positive response rates (PRRs) across doctors, nurses and allied health professionals.

## Methodology

### Study design and setting

A hospital-based descriptive cross-sectional study was conducted over 12 months at a Tertiary Care Hospital to assess patient safety culture and patient safety reporting among healthcare professionals.

### Study participants and sampling

Study participants included doctors, nurses, and allied health workers. Eligible participants were healthcare professionals with at least 6 months of work experience who were directly or indirectly involved in patient care and consented to participate. Participants were included in the study upon signing the informed consent form. Exclusion criteria included interns, students, trainees, and administrative staff with no patient contact, as well as healthcare professionals with less than 6 months of experience or not present during the data collection period.

The minimum sample size was calculated using the formula for a single population proportion, assuming a 95% confidence level, a prevalence of 50%, and a 10% margin of error, resulting in a required sample size of 96 participants. The sample was equally allocated among the three professional groups, with 32 participants each from doctors, nurses, and allied health professionals. Stratified random sampling was used, with participants first stratified according to professional group and then randomly selected within each stratum. Equal allocation was chosen to facilitate comparison of patient safety culture across the three professional groups. However, this allocation does not reflect the actual proportion of these professional groups within the hospital workforce and may therefore limit the generalizability of the findings to the overall healthcare workforce.

### Study instruments and data collection

Data collection was conducted using the Indian version of the Hospital Survey on Patient Safety Culture 2.0 (I-HSOPSC 2.0), a validated, modified version of the AHRQ Hospital Survey on Patient Safety Culture 2.0, developed specifically for India. The questionnaire was composed of a total of 32 core questions that were grouped into 10 different patient safety culture dimensions, such as teamwork, staffing and work pace, organisational learning, response to error, communication openness, communication regarding errors, supervisor/manager support, hospital management support, handoffs and information exchange, and patient safety events reporting. The questionnaire also included a section on overall patient safety rating and the occurrence of patient safety events. The Indian version of the tool has demonstrated adequate content validity (S-CVI = 0.935; I-CVI = 0.80–1.00), while the handoffs and information exchange dimensions showed low internal consistency in India and were therefore omitted from the composite score analysis, as established in the validation study. When analysing the results, the negatively worded questions were converted to positive ones per the AHRQ HSOPSC 2.0 scoring instructions.

### Statistical analysis

The data was analysed using SPSS Statistics version 30. Continuous variables are reported as means and standard deviations; categorical variables as frequencies and percentages. The patient safety culture evaluation was performed by calculating the Positive Response Rate (PRR) for each survey question, following the I-HSOPSC 2.0 user guide. The normality of scores for each domain was assessed by the Shapiro-Wilk test. Since the data was not normally distributed, the appropriate non-parametric statistical methods were utilised. Given that patient safety culture responses utilised Likert-type ordinal scales, rank-based non-parametric methods were deemed suitable for comparing outcomes among independent professional groups [15]. Differences in PRR values among doctors, nurses, and allied health professionals were examined using the Kruskal-Wallis test. For categorical variables with expected cell counts less than 5, Fisher’s exact test was used. The two-tailed p-value <0.05 was considered statistically significant.

### Ethical considerations

To conduct the study, administrative approval was obtained from the Hospital via a No Objection Certificate (NOC) prior to data collection. The study guidelines were reviewed and approved by the Institution’s Scientific Review Board. All the participants were informed about this process. Participants volunteered, and they gave their consent prior to data collection. The confidentiality of participants was ensured throughout the research process, affording them the right to opt out at any time.

## Results

The present study assessed patient safety culture among healthcare professionals, namely doctors, nurses, and allied health professionals, at a tertiary care hospital using the validated Indian version of the Hospital Survey on Patient Safety Culture (I-HSOPSC 2.0). A total of 96 healthcare professionals were included, with 32 participants from each professional group. The overall patient safety culture score was 3.72 ± 0.52 on a five-point scale, indicating a positive perception of patient safety culture. While the overall safety culture was favourable, opportunities for improvement were identified across several domains. The highest mean score was observed for Communication About Errors (4.14 ± 0.89), followed by Management Support for Patient Safety (3.96 ± 0.77) and Teamwork (3.90 ± 0.96). These domains received higher ratings than the remaining domains, indicating relative strengths in patient safety culture. Communication Openness, Reporting of Patient Safety Events, and Handoffs and Information Exchange also showed moderate scores (Table 1).

**Table 1.** Descriptive Statistics of Patient Safety Culture Domains.

| Variable | Mean (SD) | Median (IQR) | Shapiro-Wilk |  |
| --- | --- | --- | --- | --- |
|  |  |  | Test Statistic | p-value |
| Teamwork | $3.90 \pm 0.96$ | 4 (1.33) | 0.900 | <0.001 |
| Staffing | $3.20 \pm 0.72$ | 3.25 (1.25) | 0.953 | 0.014 |
| Org Learning | $3.83 \pm 0.78$ | 4 (1) | 0.915 | <0.001 |
| Response Error | 3.31 ± 0.54 | 3.5 (0.5) | 0.947 | 0.007 |
| Supervisor | 3.48 ± 0.72 | 3.33 (0.67) | 0.883 | <0.001 |
| Comm Error | 4.14 ± 0.89 | 4.33 (1.33) | 0.851 | <0.001 |
| Comm Open | 3.86 ± 0.84 | 4 (1.17) | 0.942 | 0.004 |
| Reporting | 3.87 ± 1.10 | 4 (2) | 0.866 | <0.001 |
| Management | 3.96 ± 0.77 | 3.67 (1.67) | 0.904 | <0.001 |
| Handoffs | 3.64 ± 0.99 | 3.67 (1.83) | 0.935 | 0.002 |
| Overall PSC | 3.72 ± 0.52 | 3.75 (0.84) | 0.964 | 0.047 |

Among the domains assessed, Staffing and Work Pace received the lowest mean score (3.20 ± 0.72), followed by Response to Error (3.31 ± 0.54). These findings indicate areas requiring further attention, particularly regarding workload, staffing adequacy, and the development of a non-punitive environment for reporting and addressing errors. The Shapiro-Wilk test indicated that the domain scores were not normally distributed, so non-parametric statistical methods were applied for inferential analysis, as shown in Table 1.

Significant differences (P<0.05) were observed among the three professional groups for Communication About Errors, Communication Openness, Management Support for Patient Safety, and Overall Patient Safety Culture. Allied health professionals generally reported higher mean scores than nurses across these domains. No statistically significant differences were observed among the three professional groups for Teamwork, Staffing and Work Pace, Organisational Learning–Continuous Improvement, Response to Error, Supervisor/Manager Support, Reporting of Patient Safety Events, or Handoffs and Information Exchange as shown in Table 2.

**Table 2.** Comparison of Patient Safety Culture Domains Among Professional Categories Using Kruskal–Wallis Test.

| **Variable** | **Participant** | **Mean ± SD** | **Median**<br>**(IQR)** | **Test**<br>**Statistic** | **p-**<br>**value** |
| --- | --- | --- | --- | --- | --- |
| **Teamwork** | Doctor | 4.16 ± 0.72 | 4.33(1.67) | 1.849 | 0.397 |
| | Nurse | 3.73 ± 1.14 | 4.00 (1.33) | | |
| | AHPs | 3.96 ± 0.78 | 4.00 (1.17) | | |
| **Staffing** | Doctor | 3.23 ± 0.86 | 3.25 (1.25) | 2.564 | 0.278 |
| | Nurse | 3.09 ± 0.73 | 3.00 (1.25) | | |
| | AHPs | 3.33 ± 0.60 | 3.75 (0.75) | | |
| **Org**<br>**Learning** | Doctor | 4.15 ± 0.60 | 4.00 (1.33) | 0.844 | 0.656 |
| | Nurse | 3.69 ± 0.81 | 4.00 (1.33) | | |
| | AHPs | 3.81 ± 0.82 | 4.00 (0.83) | | |
| **Response**<br>**Error** | Doctor | 3.40 ± 0.63 | 3.50 (0.50) | 0.656 | 0.720 |
| | Nurse | 3.21 ± 0.54 | 3.25 (0.50) | | |
| | AHPs | 3.40 ± 0.48 | 3.50 (0.50) | | |
| **Supervisor** | Doctor | 3.57 ± 0.44 | 3.33 (0.67) | 2.765 | 0.251 |
| | Nurse | 3.43 ± 0.81 | 3.33 (0.67) | | |
| | AHPs | 3.49 ± 0.76 | 3.66 (0.83) | | |
| **Comm**<br>**Error** | Doctor | 4.02 ± 0.80 | 4.00 (2.00) | 12.491 | 0.002* |
| | Nurse | 4.13 ± 0.94 | 4.33 (1.33) | | |
| | AHPs | 4.24 ± 0.90 | 4.33 (1.00) | | |
| **Comm Open** | Doctor | 4.02 ± 0.74 | 4.00 (1.67) | 12.915 | 0.002* |
| | Nurse | 3.63 ± 0.82 | 3.75 (1.25) | | |
| | AHPs | 4.08 ± 0.87 | 4.50 (1.25) | | |
| **Reporting** | Doctor | 4.03 ± 0.76 | 4.00 (1.50) | 1.441 | 0.487 |
|  | Nurse | 3.87 ± 1.04 | 4.00 (2.00) |  |  |
|  | AHPs | 3.76 ± 1.39 | 4.50 (2.50) |  |  |
| **Management** | Doctor | 4.02 ± 0.75 | 3.66 (1.67) | 9.189 | 0.010* |
|  | Nurse | 3.82 ± 0.78 | 3.66 (1.00) |  |  |
|  | AHPs | 4.11 ± 0.79 | 4.33 (1.33) |  |  |
| **Handoffs** | Doctor | 3.42 ± 1.18 | 3.66 (2.00) | 2.465 | 0.292 |
|  | Nurse | 3.57 ± 0.93 | 3.33 (1.67) |  |  |
|  | AHPs | 3.92 ± 0.90 | 4.00 (1.42) |  |  |
| **Overall PSC** | Doctor | 3.80 ± 0.51 | 3.71 (0.78) | 8.169 | 0.017* |
|  | Nurse | 3.62 ± 0.56 | 3.60 (0.80) |  |  |
|  | AHPs | 3.81 ± 0.47 | 4.00 (0.68) |  |  |

\*Statistically significant at $p < 0.05$

Table 3 shows the association between professional category and patient safety event reporting. Among participants who reported 3–5 events, 46.7% were doctors, 26.7% were nurses, and 26.7% were allied health professionals. Among those who reported 6–10 events, 38.5% were doctors, 26.9% were nurses, and 34.6% were allied health professionals. Of those reporting 11 or more events, 17.6% were doctors, 38.2% were nurses, and 44.1% were allied health professionals. However, no statistically significant association was found between professional category and patient safety event reporting (P=0.131).

**Table 3.**
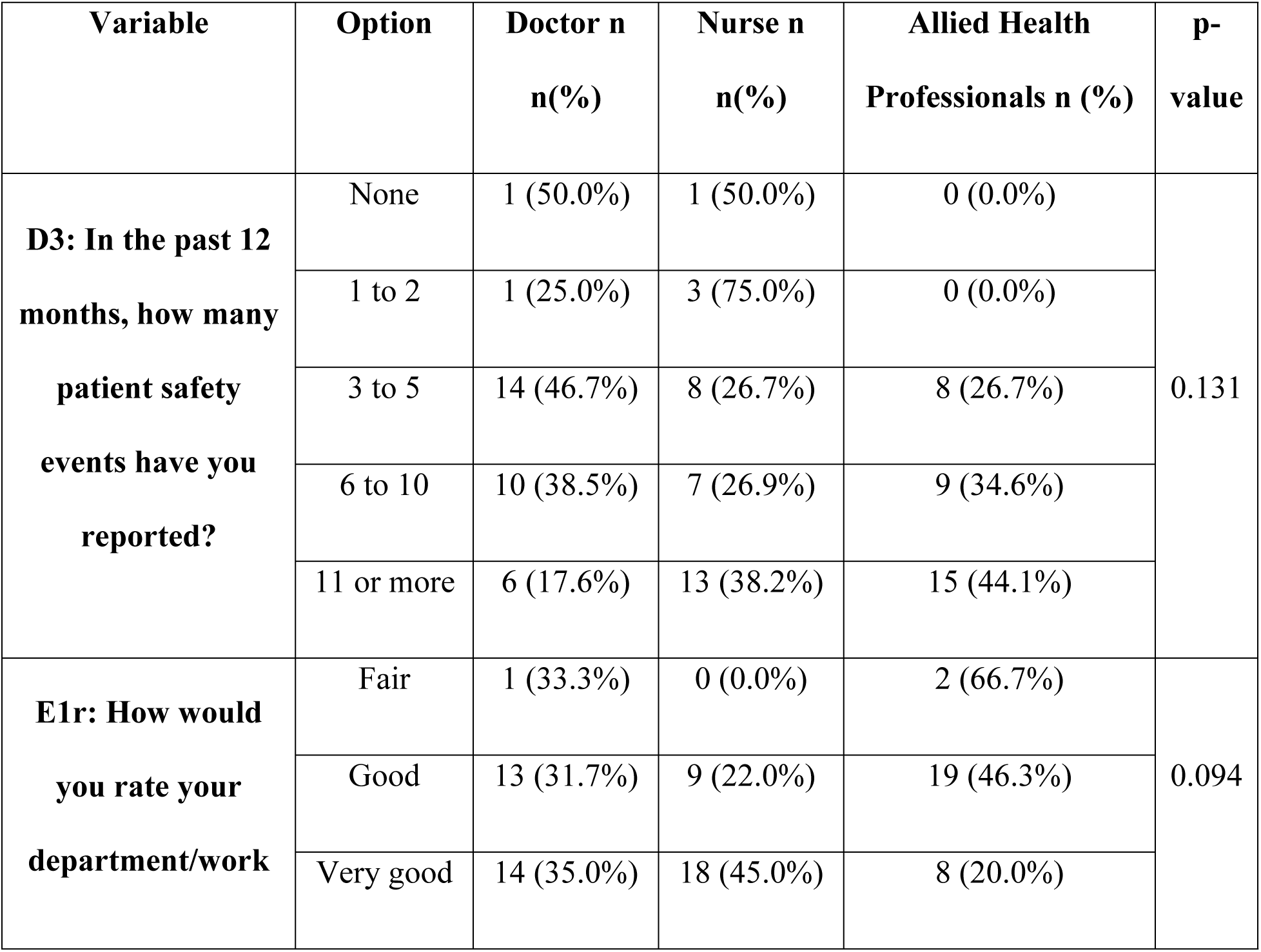
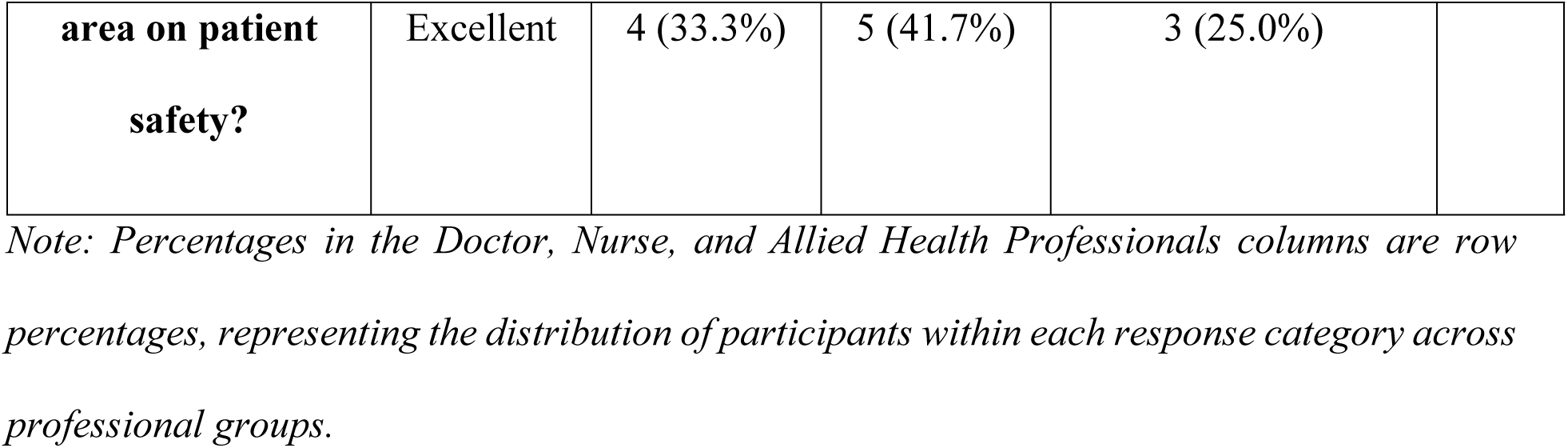
Association Between Professional Category and Patient Safety Event Reporting and Patient Safety Rating Using Fisher’s Exact Test.

The table also shows the association between professional category and overall patient safety rating. Among participants who rated patient safety as Good, 31.7% were doctors, 22.0% were nurses, and 46.3% were allied health professionals. For the Very Good rating, 35.0% were doctors, 45.0% were nurses, and 20.0% were allied health professionals, while among those who rated patient safety as Excellent, 33.3% were doctors, 41.7% were nurses, and 25.0% were allied health professionals. Although differences were observed in the distribution of ratings across professional groups, the association was not statistically significant (P=0.094).

Further indicates that the highest positive responses (PRRs) were provided mainly by allied healthcare professionals across most questionnaire items, whereas nurses tended to record the lowest PRRs. Higher PRRs correspond to the survey items related to teamwork, organisational learning, support from supervisors, communication concerning errors, and reporting safety events in a patient. In contrast, the items related to staff and working pace, open communication, and responding to errors showed low PRRs for the majority of professional groups represented in the survey. In general, the survey results show a positive culture of patient safety and high rates of reporting, but there are also opportunities to improve staffing and workload organisation, as well as openness and a non-punitive error-reporting system, as shown in Table 4.

**Table 4.** Item-wise Positive Response Rate (PRR) Among Participants.

| **Questions** | **Doctor (N=32)** | **Nurse(N=32)** | **Allied Health**<br>**Professionals(N=32)** |
| --- | --- | --- | --- |
| A1r_PRR | 96.9% | 81.3% | 93.8% |
| A2r_PRR | 78.1% | 62.5% | 53.1% |
| A3r_PRR | 6.5% | 16.1% | 25.8% |
| A4r_PRR | 75.0% | 78.1% | 81.3% |
| A5r_PRR | 30.0% | 40.7% | 53.3% |
| A6r_PRR | 51.7% | 53.1% | 45.2% |
| A7r_PRR | 63.3% | 46.9% | 71.9% |
| A8r_PRR | 100.0% | 78.1% | 90.6% |
| A9r_PRR | 24.1% | 58.6% | 75.0% |
| A10r_PRR | 10.7% | 29.0% | 9.4% |
| A11r_PRR | 40.6% | 51.7% | 71.0% |
| A12r_PRR | 81.3% | 83.3% | 84.4% |
| A13r_PRR | 55.2% | 56.7% | 75.0% |
| A14r_PRR | 64.5% | 65.6% | 87.5% |
| B1_PRR | 81.3% | 83.9% | 90.6% |
| B2_PRR | 29.0% | 28.1% | 25.8% |
| B3_PRR | 78.1% | 81.3% | 90.3% |
| C1r_PRR | 64.5% | 71.0% | 84.4% |
| C2r_PRR | 64.5% | 75.0% | 96.9% |
| C3r_PRR | 67.7% | 71.9% | 93.5% |
| C4r_PRR | 71.9% | 74.2% | 87.5% |
| C5r_PRR | 71.0% | 66.7% | 83.3% |
| C6r_PRR | 70.0% | 63.3% | 81.0% |
| C7r_PRR | 28.1% | 46.9% | 77.4% |
| D1_PRR | 54.8% | 68.8% | 80.0% |
| D2_PRR | 68.8% | 73.3% | 73.3% |
| F1_PRR | 81.3% | 81.3% | 93.8% |
| F2_PRR | 78.1% | 81.3% | 84.4% |
| F3_PRR | 28.1% | 28.6% | 71.0% |
| F4_PRR | 47.1% | 48.3% | 81.0% |
| F5_PRR | 58.8% | 67.7% | 81.8% |
| F6_PRR | 64.3% | 51.6% | 70.0% |

*Note: Item-code legend: A1r–A4r = Teamwork; A5r–A10r = Staffing and Work Pace; A11r–A12r = Organisational Learning; A13r–A14r = Response to Error; B1–B3 = Supervisor/Manager Support; C1r–C7r = Communication About Errors; D1–D2 = Communication Openness; F1–F6 = Reporting of Patient Safety Events. The “r” suffix denotes the reverse-worded item identifier used in the I-HSOPSC 2.0 scoring convention*

## Discussion

The present study found an overall patient safety culture score of 3.72 ± 0.52 among doctors, nurses, and allied health professionals, indicating a positive perception of patient safety culture. This indicates that the hospital has implemented numerous favourable practices for patient safety, but certain improvements are still needed. The highest assessment was given to communication regarding errors, administrative support for patient safety, and teamwork dimensions among those examined in I-HSOPSC 2.0. The findings align with the observations of Aileen J. et al. and Vijay K. Tadia et al. regarding the collective nature of health care professionals’ work and communication about patient safety activities, which is a strong point in patient safety culture in India [1,2].

Even though there were positive outcomes, the pace of work and handling of mistakes were seen as the weakest parameters. In other words, healthcare workers still must deal with organisational issues. Those issues revolve around insufficient staffing, heavy workloads, and the handling of mistakes. Consequently, these factors lead to fatigue, poor communication, and an unwillingness to report incidents, which eventually undermine patient safety. Akoijam et al. also reported similar concerns regarding staffing and work pace in an Indian study conducted among nurses at two tertiary-care hospitals in Manipur [12]. Similar findings were reported by researchers Livia Margarita et al. and Gelana Fekadu et al., who claim that insufficient staffing, limited workplace resources, and a punitive approach to mistakes are major impediments to establishing a proper culture of patient safety [6,3].

The current research also revealed differences in perceptions of patient safety culture among professional groups. Allied Health professionals in general reported a more positive feeling about patient safety than nurses across many dimensions, while nurses showed relatively the least positive responses in comparison to other professions. The reason may be related to differences in responsibilities and workload. Nurses typically have greater responsibility for continuous patient monitoring, completing documentation, and collaborating with other doctors and professionals, which may influence their attitudes toward staffing adequacy and workplace safety. Similar differences among professional groups were found by Annamaria Bagnasco et al., showing that views on patient safety culture depend on roles and responsibilities within the organisation [4]. A recent study by Marathe et al., conducted in a public tertiary-care teaching hospital in Central India, examined patient safety culture among doctors, nurses, and other healthcare professionals. The study reported significant differences in perceptions across these professional groups [13].

While most participants reported documenting multiple patient safety incidents in the previous year, many still viewed their departments as safe. This indicates that awareness of the importance of incident reporting has increased, but there is still room for improvement and a need to strengthen reporting systems and the organisational response. Previous studies from Vijay K. Tadia et al., Livia Margarita et al., and Gelana Fekadu et al. found similar barriers to effective patient safety event reporting, namely blame-related fears, lack of reporting systems, and lack of organisational support [2,6,3].

Overall, healthcare professionals had a positive perception of patient safety culture, with teamwork, communication about errors, and management support identified as key strengths. Nonetheless, the adequacy of staffing and work pace, along with the organisation’s response to errors, continues to stall the establishment of a more mature safety culture. Hence, even though the institution has a solid basis for improving patient safety, there should be sustained efforts to eradicate understaffing, create a non-punitive environment for error reporting, and provide greater organisational support. Implementing the necessary measures in these areas would both improve the patient safety culture and improve the quality of work.

### Limitations

This study has several limitations. First, the findings were obtained from a single tertiary care hospital and therefore may not be generalizable to primary, secondary, or other public healthcare institutions. Second, the sample size of 96 participants was relatively small and, although equally distributed among doctors, nurses, and allied health professionals, may not fully represent the wider population of healthcare workers. Third, the use of a self-reported questionnaire may have introduced social desirability bias, as participants may have provided responses they perceived as more favourable rather than reflecting their actual practices or experiences. Fourth, the cross-sectional design limits the ability to assess changes in perceptions of patient safety culture over time. Finally, the relatively high proportion of “Does not apply/Don’t know” responses for some items related to Management Support and Communication may have affected the interpretation of these items and should be considered when evaluating the findings.

## Conclusion

The study found a positive perception of patient safety culture among healthcare professionals. Communication about errors, management support for patient safety, and teamwork were the strongest domains, whereas staffing, work pace, and response to error were priority areas for improvement. Differences among professional groups, particularly the comparatively lower scores among nurses in several domains, support the need for profession-specific interventions. These findings highlight the need for hospital-level strategies that address staffing and workload concerns while strengthening communication, error reporting, and management support for patient safety. Strengthening workforce capacity, promoting a just and non-punitive approach to error reporting, and improving communication can support continued development of patient safety culture.

## Data Availability

The coded data set will be made available

## Acknowledgment

The authors would like to express their sincere gratitude to the management and healthcare professionals at the participating hospital for their support and cooperation throughout this study. The authors also acknowledge the support and guidance provided by the Institutional Scientific Review Board.

## Data Availability Statement

The data underlying the findings of this study cannot be made publicly available due to confidentiality requirements stipulated by the participating hospital. The hospital’s permission specified that all data collected was to be used strictly for academic and research purposes and maintained confidentially in accordance with hospital policies. Therefore, the underlying data cannot be shared publicly.

## Ethics Statement

The research proposal was reviewed and accepted by the Institutional Scientific Review Board. Permission to conduct the study was also obtained from the participating hospital.

## References

1. Aileen J, Pushpanjali K, Federico F, Joseph L, Manjunath U. Psychometric analysis of the Indian version of the Patient Safety Culture Tool (I-HSOPSC 2.0) validation. J Health Manag. 2024;26(1):109–115. doi:10.1177/09720634231216564.

2. Tadia VK, Kotwal N, Jalaunia RS. Patient safety culture: insights from a cross-sectional study among healthcare professionals. J Family Med Prim Care. 2025;14:90–96. doi:10.4103/jfmpc.jfmpc_904_24.

3. Fekadu G, Muir R, Tobiano G, Bime AE, Ireland MJ, Marshall AP. Patient safety culture in resource-limited healthcare settings: a multicentre survey. PLoS One. 2025;20:e0326320. doi:10.1371/journal.pone.0326320.

4. Bagnasco A, Catania G, Loiudice MT, Bellandi T, Cavaliere B, Carzaniga S, et al. Validation of the Hospital Survey on Patient Safety Culture 2.0 in Italian hospitals: a cross-sectional study of healthcare personnel perceptions. J Adv Nurs. 2025;81:7609–7632. doi:10.1111/jan.16770.

5. Wu Y, Hua W, Zhu D, Onishi R, Yang Y, Hasegawa T. Cross-cultural adaptation and validation of the Chinese version of the revised surveys on patient safety culture (SOPS) hospital survey 2.0. BMC Nurs. 2022;21:369. doi:10.1186/s12912-022-01142-3.

6. Margarita L, Chalidyanto D, Tanaya EM. Assessment of patient safety culture at a private hospital in Surabaya: a cross-sectional study using HSOPSC 2.0. STRADA J Ilm Kesehat. 2025;14(2):216–225. doi:10.30994/sjik.v14i2.1226.

7. Chen IC, Li HH. Measuring patient safety culture in Taiwan using the Hospital Survey on Patient Safety Culture (HSOPSC). BMC Health Serv Res. 2010;10:152. doi:10.1186/1472-6963-10-152.

8. Rajalatchumi A, Ravikumar TS, Muruganandham K, Thulasingam M, Selvaraj K, Reddy MM, et al. Perception of patient safety culture among health-care providers in a tertiary care hospital, South India. J Nat Sci Biol Med. 2018;9:14–18. doi:10.4103/jnsbm.JNSBM_86_17.

9. Azyabi A, Karwowski W, Davahli MR. Assessing patient safety culture in hospital settings. Int J Environ Res Public Health. 2021;18:2466. doi:10.3390/ijerph18052466.

10. Imran Ho DS, Jaafar MH, Mohammed Nawi A. Revised Hospital Survey on Patient Safety Culture (HSOPSC 2.0): cultural adaptation, validity and reliability of the Malay version. BMC Health Serv Res. 2024;24:1287. doi:10.1186/s12913-024-11802-6.

11. Bhaladhare R, Rishipathak P. A Cross-Sectional Study on Patient Safety Culture in a Tertiary Care Hospital in India. Int J Stat Med Res. 2025;14:920–928. doi:10.6000/1929-6029.2025.14.81.

12. Akoijam P, Konjengbam S. The impact of an educational program on knowledge and perception of patient safety culture among nurses in the two medical colleges of Manipur: a quasi-experimental study. Indian J Public Health. 2023;67(2):265–270. doi:10.4103/ijph.ijph_1416_22.

13. Marathe N, Varghese A, Patil D, George G, Quadri SR, Singh V, et al. Patient safety culture in a public tertiary care teaching hospital in Central India: a cross-sectional study using the Hospital Survey on Patient Safety Culture (HSOPSC) Version 2.0. Cureus. 2026;18(3):e105740. doi:10.7759/cureus.105740.

14. Khalek MA, Huq KATM E, Hasegawa T, Hatakeyama Y, Babaita AO, Moriyama M. Cross-cultural adaptation and validation of hospital survey on patient safety culture in Bengali version (B-HSOPSC 2.0) among nurses in Bangladesh: a cross-sectional study. BMC Nurs. 2026;25:679. doi:10.1186/s12912-026-04817-3.

15. Al-Jaishi AA, Cuerden MS, Luo B, Roshanov PS, Garg AX. Statistical analysis of Likert-based ordinal scales: a guide for clinical trialists. BMC Medical Research Methodology. 2026;26:78. doi:10.1186/s12874-026-02793-5.

